# Farr’s Law and Risk Aversion in the Study of Epidemics

**DOI:** 10.1101/2025.04.01.25325028

**Authors:** Michael Brimacombe

## Abstract

The modeling of the frequency of new cases of infection in an epidemic within a given time period remains of great interest. Farr’s Law and the related Brownlee-Farr condition on infection counts implies a specific functional form for the relative infection incidence rate *I*(*t*) over the course of the epidemic. This form includes the Gaussian distribution, but also other distributions such as the exponential distribution. The Brownlee-Farr condition is shown to correspond as a mathematical function to the Pratt-Arrow ARA measure of relative risk avoidance, and also the hazard rate, with differences due to slight variations in the chosen units of measurement. These types of linkages may allow for the development of new areas of application for the various underlying model types, implying a greater flexibility in the choice of statistical model.

## 1 Introduction

Epidemics of infectious disease in human and animal populations have been the focus of study for many years. The onset, intensity and duration of such outbreaks have often been well documented, with the reasons for each of these being the source of much study [1]. Once an epidemic is ongoing, it is important to understand whether efforts to mitigate the spread of the epidemic are working. Often, an important observation in relation to an epidemic weakening is that the rate of increase in the number of cases of infection should be decreasing as the epidemic moves forward in time [1].

In the past, without modern medical treatments and vaccinations, the onset of measles, smallpox, polio, pertusis, cholera, typhoid, malaria, tuberculosis and other infectious threats to health were a constant threat. Many of these still exist, and without serious vaccination campaigns, remain a threat [2].

Formal mathematical models of epidemics [3] provide important insights. For example, the ability to model the relative infection incidence rate *I*(*t*) as a function of time allows government and public health officials to predict possible disease related scenarios and better prepare needed resources. Often cases are aggregated into weekly or monthly counts and these are standardized in various ways to provide a data based picture of the epidemic, usually in the form of probability distributions and related predictions.

There is a history of using probability distributions and stochastic models to summarize infection-related data patterns through time [4]. There are various distributions that can be fit or trained to existing data. These include the binomial, poisson, exponential and normal distributions usually in the context of time series data or stochastic processes. They may also provide useful predictions regarding these patterns [5]. Note that central limit theorems related to statistical frequentist sampling theory may also apply in these often large-sample settings, affecting probability based calculations.

In regard to the modeling of incidence rates in the specific context of epidemics, early work by Farr [6] showed that a Gaussian distribution, written as a function of time, often arises when the pattern of relative incidence *I*(*t*) obeys a specific condition in the context of epidemics. This was further developed by Brownlee [7, 8]. Expressed as a function of time, if this Brownlee-Farr (BF) condition is satisfied, the onset of new cases will decrease, the frequency of infection will reach a maximum and decrease towards zero over time. This result has also been shown to hold when assuming compartmental models for *I*(*t*), sets of differential equations linking *I*(*t*) to specific mechanisms describing how the infection spreads at the level of individual interactions. For example, the SIR model [9] and the IDEA model [10, 11, 12].

Here, in the context of an ongoing epidemic or pandemic, it is shown that the BF condition may be considered from a different perspective, linking the obtained form of the *I*(*t*) function to mathematical structures that reflect the assumption of mandated wide-spread risk avoidance behavior in the general population. Such risk avoidance behavior can be modeled using the Pratt-Arrow absolute risk aversion (ARA) measure of relative risk [13] and this is shown to have a striking mathematical similarity to the BF condition. It is also shown that the hazard rate [14], defined in relation to the *I*(*t*) distribution, can also be expressed in a manner related to the ARA measure and BF distribution. In fact, the Brownlee-Farr condition and related distribution, the ARA measure and the hazard rate can be viewed as closely related measures, using different scales of measurement. The R statistical package (version 4.1.2) was used for all calculations and graphics.

## 2 Defining the Brownlee-Farr condition

Beginning with Farr’s earlier work [6] observing that the relative number of cases in an epidemic often follows an approximate Gaussian or normal distribution, Brownlee [7, 8] developed a more formal mathematical argument to explain the observation.

Brownlee argued that the loss of infecting power of an infectious agent might occur in primarily three ways, (1) the form of the curve of the epidemic, (2) the variation in the infectivity of the organisms of certain diseases between epidemic and inter-epidemic times, and (3) the nature of the spatial distribution of the epidemic, often having a well-marked centre of great prevalence, from which the amount of disease radially decreases. Note that acquired immunity is also a possibile factor, though with a newer, quickly mutating infection, this might be limited.

The argument developed by Brownlee [7, 8] assumes that at time *t* there are *y* cases. After a unit of time equal to the incubation period, there would be *y* + *dy* cases and (*y* + *dy*)*/y* is the relative “infectivity” at time *t*. Assuming this is a function of time, this can be expressed:

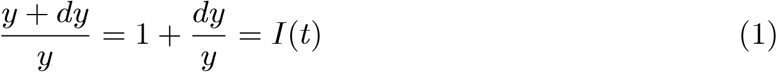

Assuming *I*(*t*) is approximately continuous, and the equation holds for small values of *t*, say *dt*, it is assumed that:

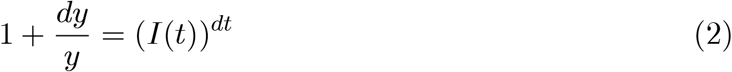

Taking the logarithm of both sides and using the Taylor series based approximation log(1 + *x*) = *x* this can be re-expressed as:

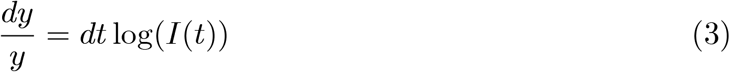

or,

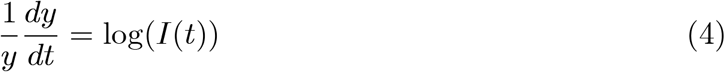

implying that the relative increase in cases, in relation to a small change in time, is given by log(*I*(*t*)).

Assuming infectivity decreases over time, *I*(*t*) is further assumed to follow a geometric progression with *I*(*t*) = exp(*a* − *bt*). Solving the resulting equation:

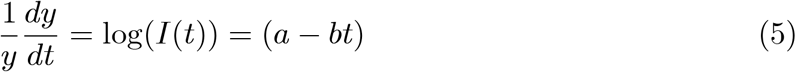

gives the relative number of cases *y*:

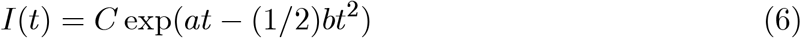

where *C* is a constant and *t* ∈ (0, ∞). This general form, once standardized, gives the normal distribution with parameters *a* and *b* to be fitted to the observed data and is denoted *I*(*t*). For specific choices of *a* and *b* this may also support several other distributions. Brownlee [7, 8] also examines more complex assumptions regarding the frequency distribution of infection.

Note that the assumption of an exponential geometric progression function implies that the first derivative of *I*(*t*) is proportional to the second derivative and their ratio is a constant, the BF condition.

## 3 Applying the Brownlee-Farr Condition

The pandemic or epidemic setting provides an interesting case study of how time-indexed relative count data values can approximately follow a normal density, when subject to simple restrictions. These patterns may arise multiple times if the infection is, for example, subject to seasonal effects [15]. The assessment of the practical application of Farr’s law might be restricted to data counts obtained from a specific area and time period, as well as, in some cases, stratifying on age group.

The BF “law”, expressed in its simplest form for a discrete setting, shows that an approximate bell-curve shape can describe the pattern of incidence at time *t, I*(*t*), for a designated time period.

If *I*(*t*) is subject to the restriction:

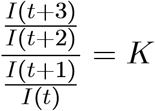

where *K* is a constant, then the incidence rate in the epidemic will occur in a manner described by a function of the general exponential form:

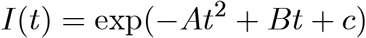

as developed in Brownlee [7, 8]. Note that the BF can be interpreted as first and second order percent differences in the time series of monthly infection counts, a ratio of re-scaled acceleration to initial percent difference.

Practically, when the Gaussian distribution does arise, the parametrized relative counts *I*(*t*) may be directly modeled as:

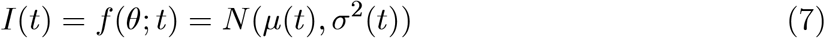

with absolute counts given by *n* · *f* (*θ*; *t*) at the observed values of *t*. Note this is only approximately a Gaussian distribution as *I*(*t*) is usually a discrete count (daily, weekly or monthly) and the normal curve an idealized approximation. See Figure 1.

**Figure 1:**
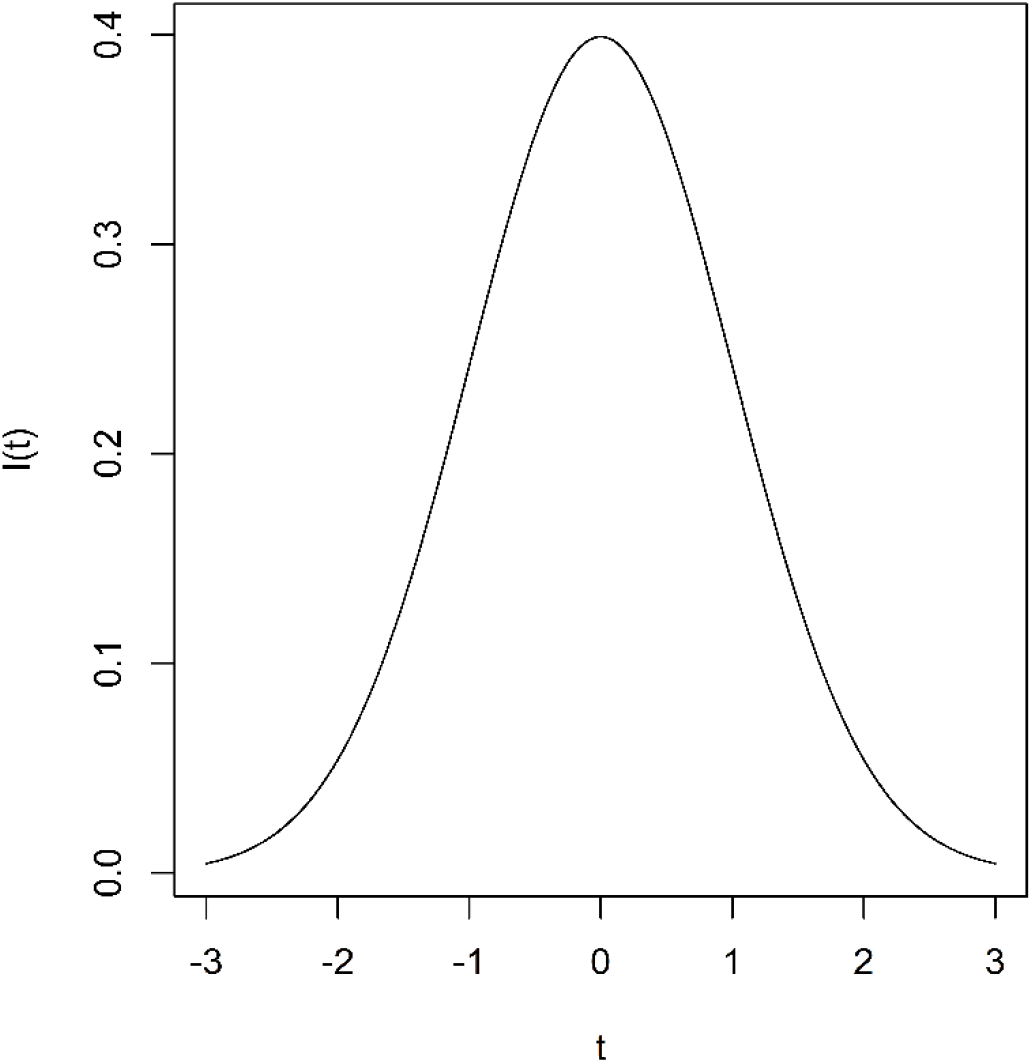
Standardized Normal Distribution.

## 4 Cattle Epidemic

The BF restriction, often most applicable within a chosen time period, has been observed in several epidemic settings [16, 17]. For example, as noted in [8], William Farr examined observed counts of cattle infected with cattle plague in England during 1865-66. He was able to develop predictions for monthly counts. These estimated values were then compared to the observed values. The results are shown in Table 1.

**Table 1:**
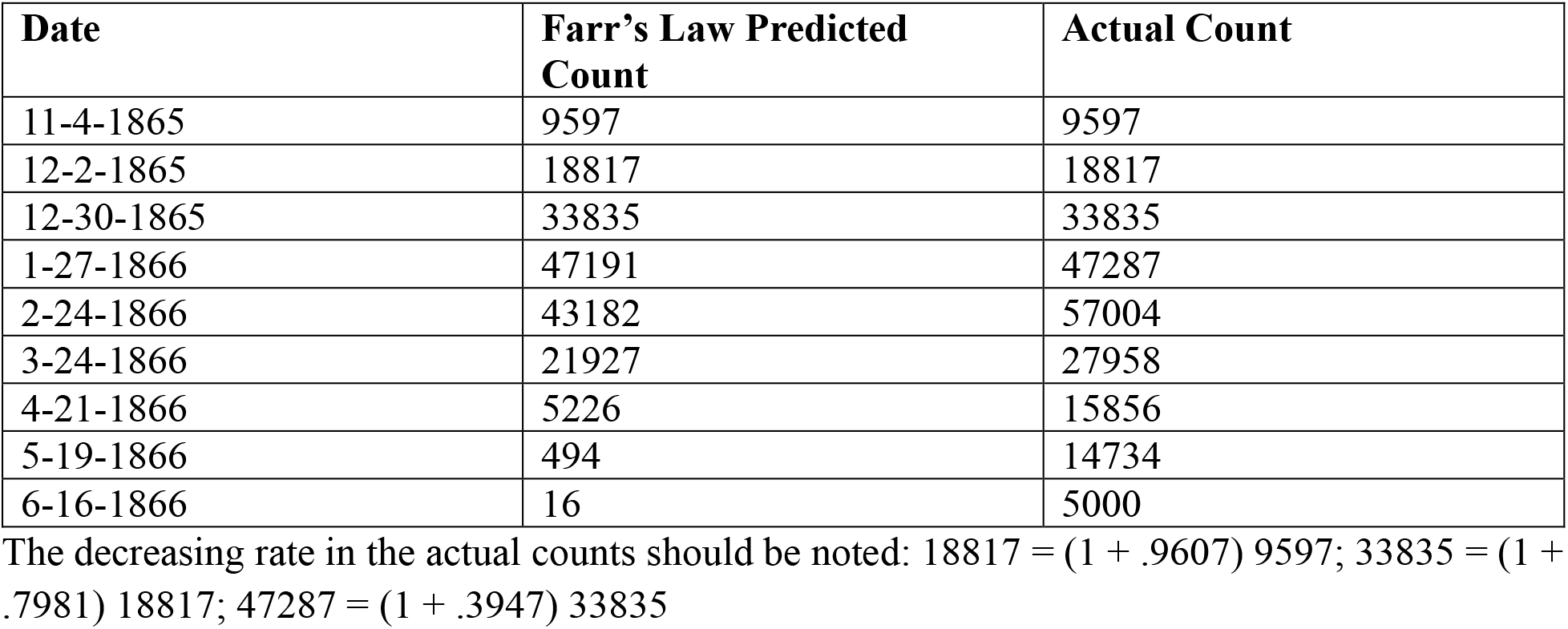
Actual and Estimated Cattle Plague Data. **Cattle Plague Data (Brownlee 1915)**

Of interest here is the decrease in the acceleration of the monthly counts. This is a key aspect in the subsequent decrease in cases and finally, an end to the epidemic. The Gaussian distribution provides an accurate description of the pattern of monthly infections. See Figure 2.

**Figure 2:**
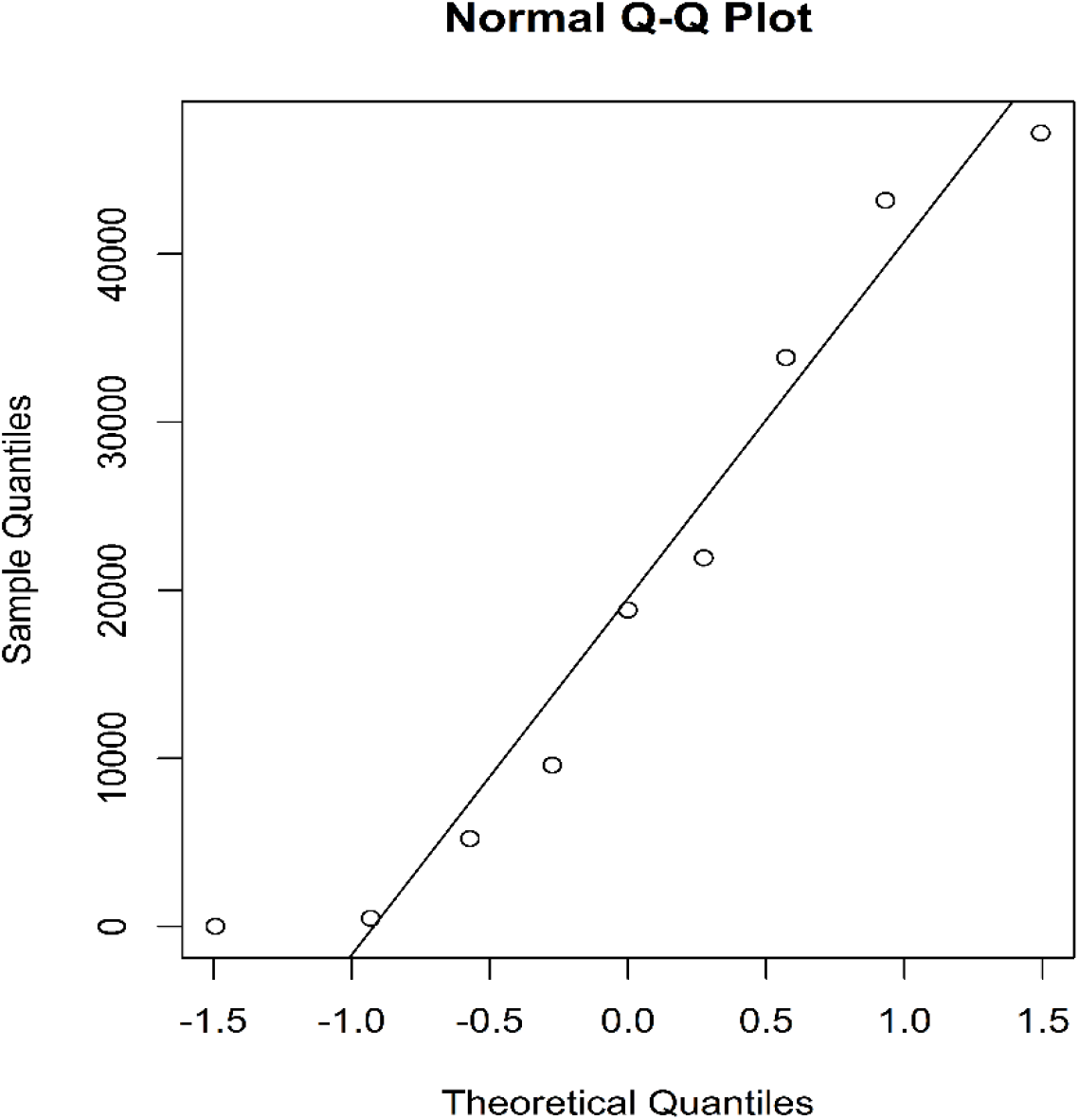
Normality Plot for Farr Estimated Values.

Note that the values estimated by Farr are conservative, but capture the maximum values of the epidemic fairly well.

### 4.1 COVID-19

To see this type of summary pattern in relation to the recent COVID-19 pandemic, the Gaussian distribution can be seen fitting the *I*(*t*) functions for a variety of countries for the period 2020 - 2022 [18]. The *I*(*t*) functions defined across time periods and countries in the early stages of the pandemic do not show stable patterns. However, over time, as societal level risk avoidance behaviors, including government restrictions and vaccines, were applied in several ways, the *I*(*t*) function stabilized into an approximately Gaussian distribution corresponding to the onset of the BF condition. See, for example, plots of COVID infections for the time period 2020 - 2023 [18].

### 4.2 Measles Epidemics

Measles is much more common than COVID-19 and has a long history. Measles epidemics have reflected the same basic BF patterns in the past [19], but are currently less reflective of epidemic related behavior as the onset of measles is limited through ongoing vaccination programs [20]. Such programs allow for greater control of the infection and this is shown in a stable distribution for *I*(*t*) which, in many countries, is close to a uniform distribution for long periods of time with only few serious outbreaks and epidemic-like behavior. See for example [21]. Figure 3 shows a plot of total measles infection reported in Afghanistan for the years 1991 - 2013. The overall median line is shown.

**Figure 3:**
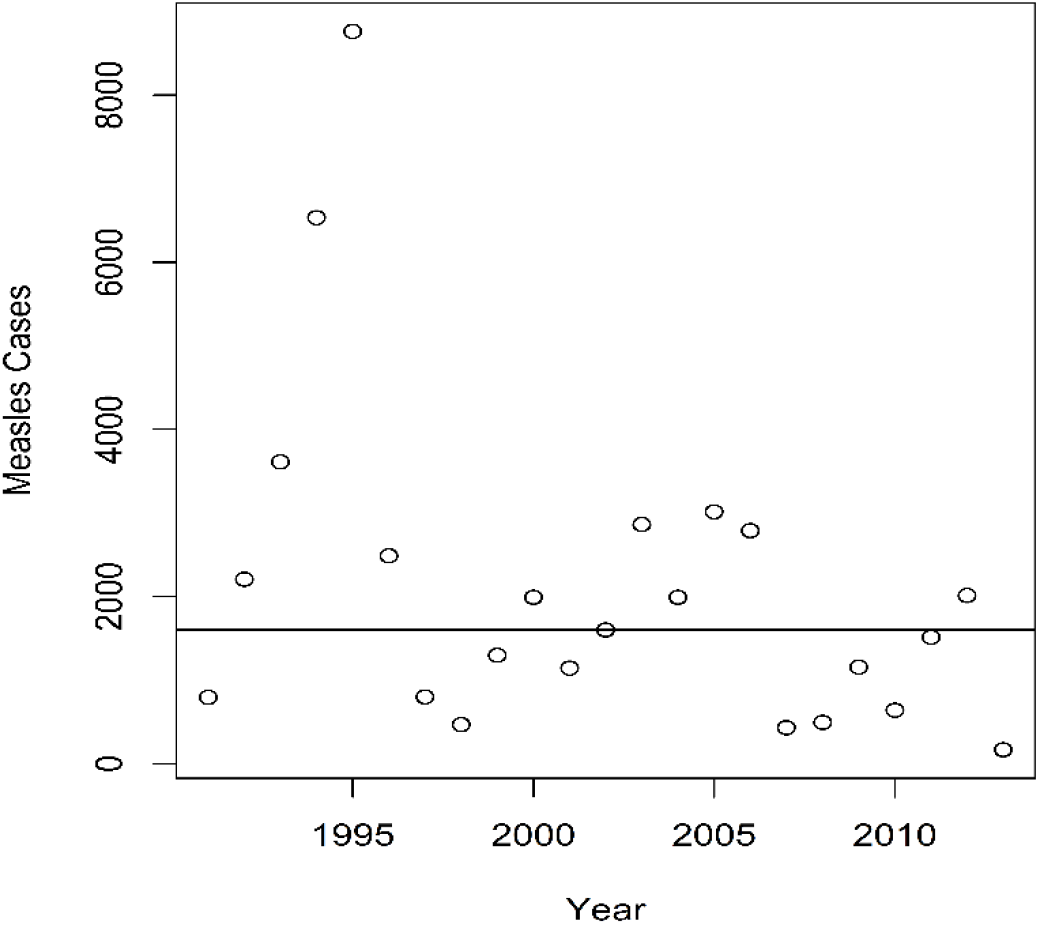
Plot of Yearly Measles Data (Afghanistan 1991 - 2013)

## 5 Probability Distributions

Assuming *I*(*t*) can be modeled directly using a probability distribution with parameter *0*, the BF condition can be examined:

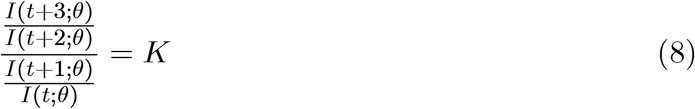

Typically the distribution *I*(*t*; *θ*) is discrete, for example monthly counts, but can be approximated using continuous distributions.

Many of the distributions related to the modeling of epidemics, such as the binomial, geometric, gamma, Poisson and Normal distributions are members of the statistical exponential family of distributions. These can be expressed in the exponential family canonical form for the single parameter *θ*:

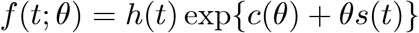

where *h*(*t*) is a norming constant, *c*(*θ*) a function of *θ* and *s*(*t*) sufficient statistic. Not all such distributions satisfy the BF condition.

Note that large sample Gaussian approximations may apply to standardized versions of these distributions [22] and are typically applied in relation to the standardized maximum likelihood estimator 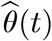 for the parameter *θ*:

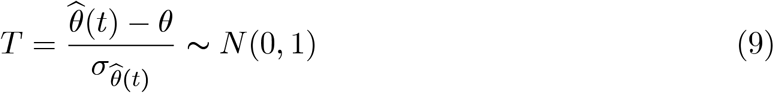

Such large-sample arguments can be applied in the context of epidemics to stochastic processes, often monthly counts, having such distributions and can reflect various assumptions. See for example Feller [23].

### 5.1 Uniform Distribution

The uniform distribution defined across a specific time period, *I*(*t*) = *k, t* ∈ [*t*_0_, *t*_l_] trivially satisfies the BF condition. An incidence function *I*(*t*) having a uniform distribution is typically not used to describe pandemic situations. It often reflects settings where the infection in question is limited. See Figure 4.

**Figure 4:**
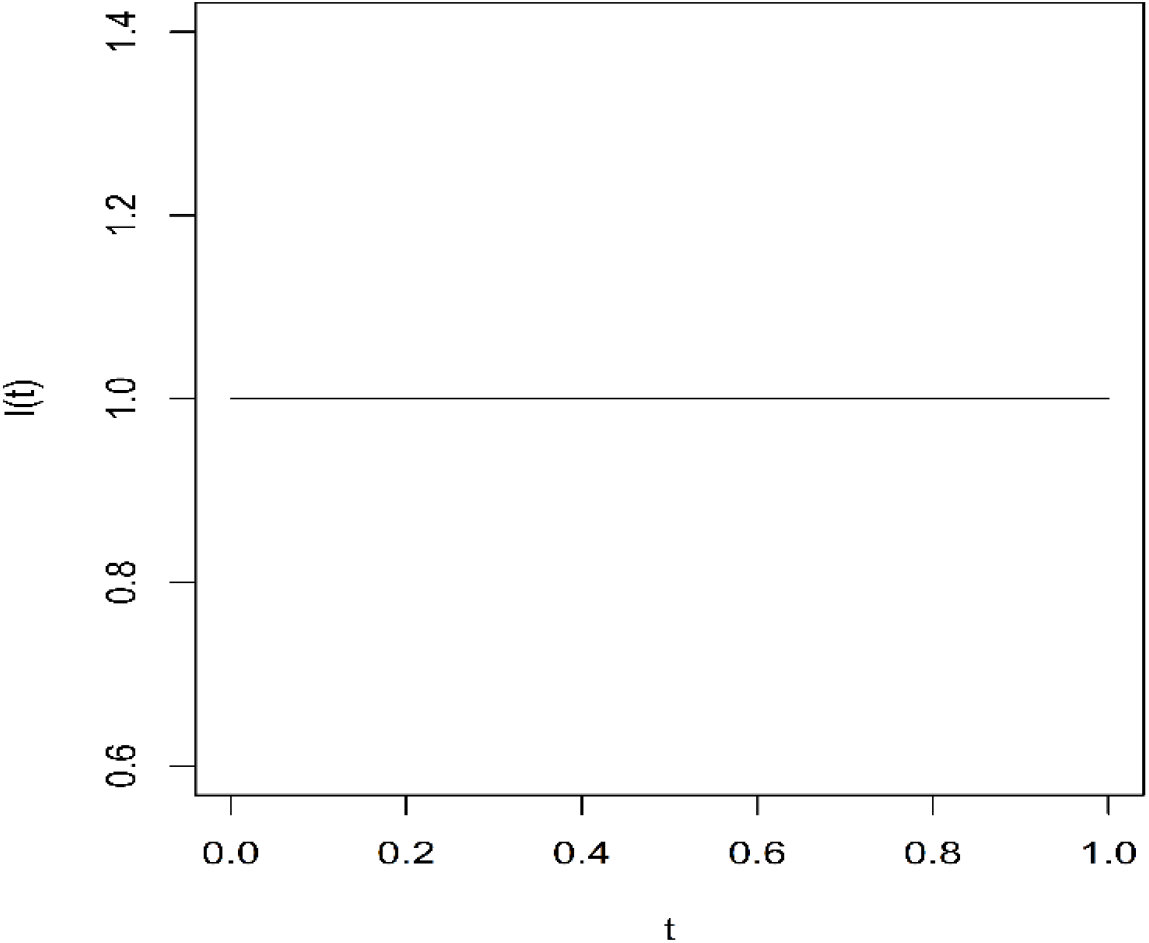
Uniform Distribution.

### 5.2 Exponential Distribution

Consider the exponential distribution which can be written:

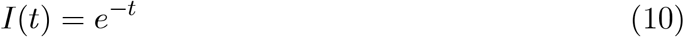

where *t* > 0. The BF condition here is given by:

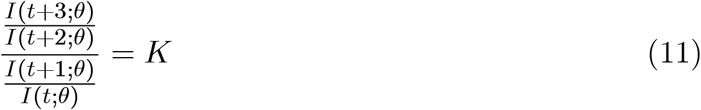

and basic algebra shows this to be true.

A more flexible format is:

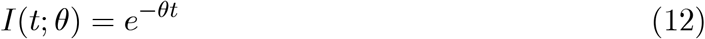

and this also satisfies the BF condition. See Figure 5.

**Figure 5:**
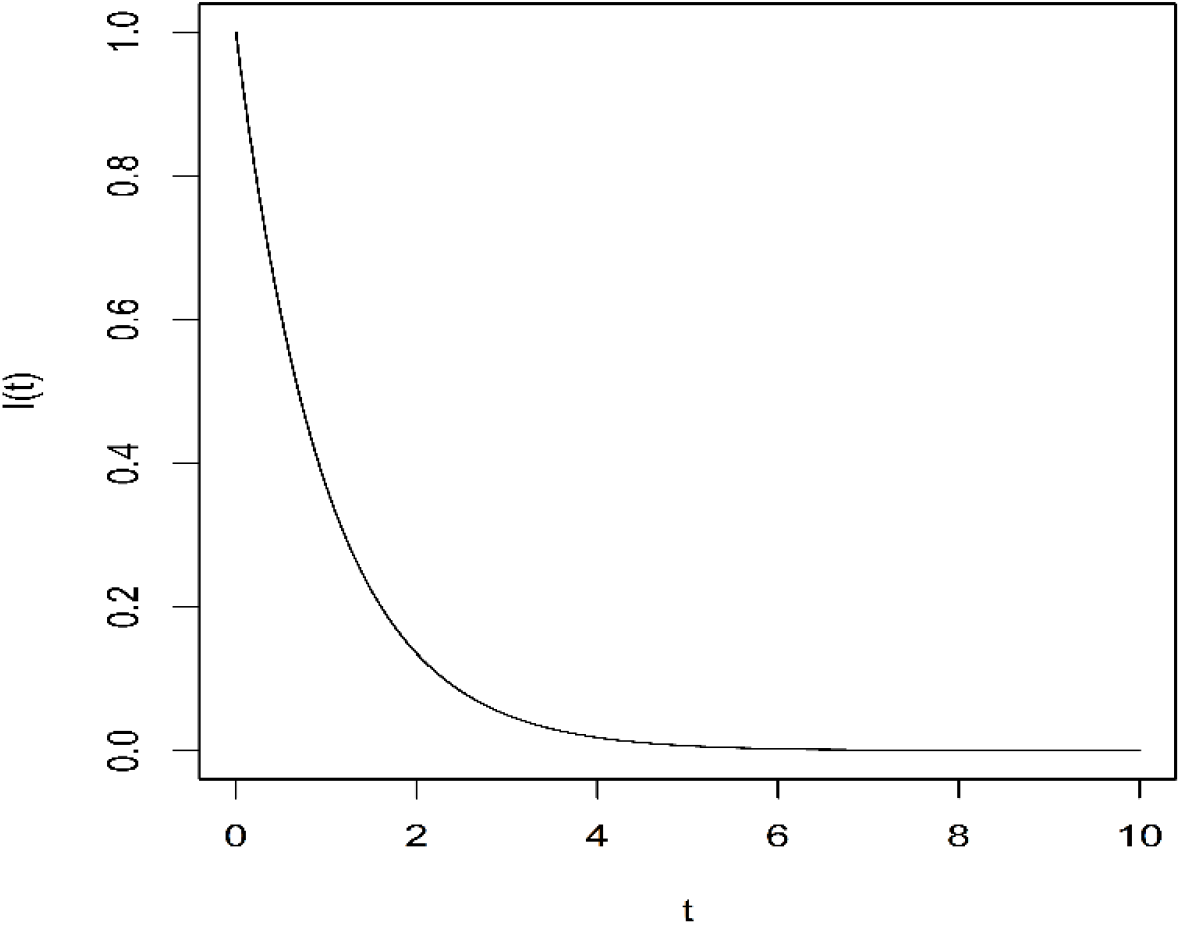
Exponential Distribution.

### 5.3 Poisson Distribution

The Poisson distribution is commonly used to model case counts that occur within a given time period. The Poisson postulates can be used to guide its application [24], though the possibly correlated nature of pandemic counts across subsequent time periods may limit its usefulness. Formally *θ* > 0, *I*(0) = 0 and the number of events in any interval of length *t* is a Poisson random variable with mean *θt* where *θ* is a constant.

Here this is used as a density formula to be fitted to the actual counts *I*(*t*). In general, a Poisson counting process can be written:

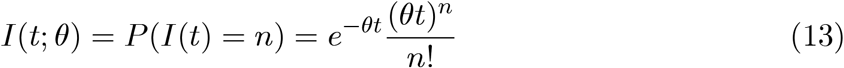

The BF condition is given by:

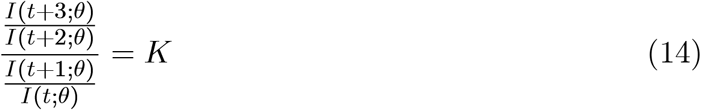

and it follows that:

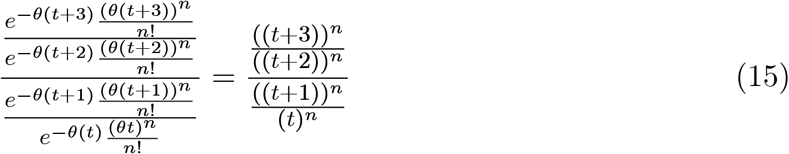

which remains a function of *t*, not satisfying the BF condition.

## 6 Large Sample Approximations

The use and interpretation of frequentist sampling distributions in relation to *I*(*t*) and the BF condition, for example the exponential family of distributions, requires caveats regarding large sample effects, including central limit theorems.

For example, note that as *n* → ∞, the Poisson distribution takes on an approximate Gaussian *N* (0, 1) density once standardized. This version then technically satisfies the BF condition. The central limit theorem here then takes the form:

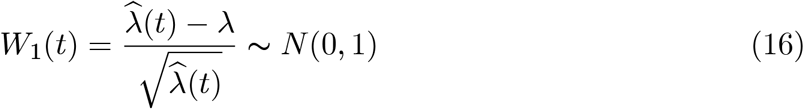

and the standardized incidence function *I*(*t*) can be expressed as:

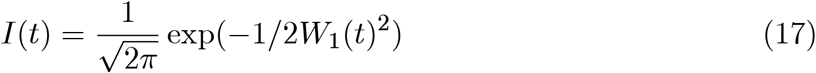

Therefore, with large samples, as discussed below, a social context supporting the onset of risk avoidance behavior or other effects on infectivity should be present when claiming the BF condition is related to the onset of a Gaussian distribution. Other distributions such as the gamma and Weibull distributions, once standardized, can also be used to model incidence data in the form of monthly counts and, in large samples, will provide sampling distributions that are approximately Gaussian. This again does not necessarily imply the BF condition holds in such cases for the underlying distributions in large samples.

## 7 Individual Micro-level Infection Models

Interestingly, the BF condition and result, as it includes functions that are not probability densities, based on macro-level considerations, can be related to compartmental models reflecting individual-level infection processes, such as the IDEA model which is based on differential equations [8]. As the IDEA model can be algebraically related to the SIR model, the BF result can be extended as well to the SIR model [9].

For example, the basic structure of the IDEA model effectively assumes:

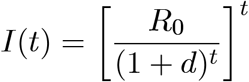

where *d* is a constant and *R*_0_ reflects the probability of transmission of the infection at the individual level. It can be shown that the BF condition holds for this functional form of *I*(*t*) giving:

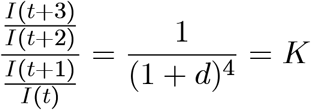

As *R*_0_ increases, the number of infected increases and the threat to healthy individuals increases. Note that while the *R*_0_ value is conceptually useful to consider when modeling the spread of an infection at the individual level, it does not formally appear in the maximization here for the version of the IDEA model given.

The *I*(*t*) function, assuming the IDEA model and thus the BF condition holds, can be written:

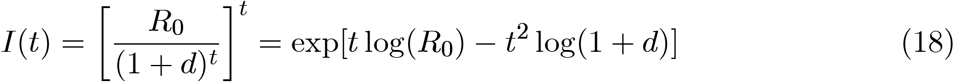

giving the BF result *I*(*t*) = exp(−*At*^2^ + *Bt* + *c*) with *A* = log(1 + *d*), *B* = log(*R*_0_), *C* = 0.

## 8 Interpretation in Relation to Macro-Level Societal Risk

The general form of the BF restriction is of interest. Practically, it is a function of time and the incidence of existing infection *I*(*t*), which itself can be viewed as a risk factor for those not yet infected. This risk will initially increase over time as the pandemic grows, modulating and weakening as the pandemic fades and this is reflected in the Gaussian-type shape of *I*(*t*) satisfying the BF condition.

In this way the a possible interpretation for the BF condition is that it is reflecting adjustments to behavior in the population, where the number of infected cases, usually counted in a specific spatial region, is perceived as representing increasing social risk in relation to the healthy, risk adverse general population.

While the BF model and restriction does not model individual level infection dynamics of the nation or community in question, leaving out, for example, an *R*0 value reflecting the speed of infection spread at the individual-to-individual level, it can usefully be interpreted as reflecting enforced common behavioral patterns, often in the form of government mandates, in the overall community and how these affect the pattern of infectivity of the epidemic over time.

## 9 The Brownlee-Farr Condition: Connections

In order to compare the BF condition and related distribution to the concepts of risk avoidance and the onset of infection defined by the hazard rate, the scale on which changes in the level of infection are measured need to be carefully considered.

The BF condition is expressed in terms of a stochastic process *I*(*t*) where differences are often represented by differences between lagged values of the sequence.

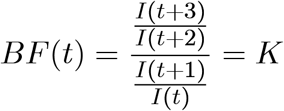

In this setting, the first-order derivative (essentially the time related relative difference or derivative Δ*I/*Δ*t*) can be written:

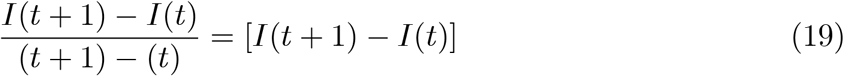

and related second-order changes:

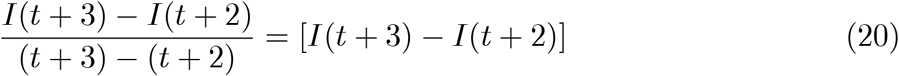

where the second-order difference in the BF condition was chosen to not overlap time points.

The BF condition itself sets a ratio of such second-order to first-order percent changes on a logarithmic scale equal to a constant. Taking a log transformation of both sides of the BF condition gives:

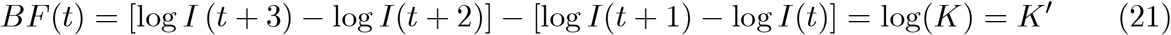

This is the difference of first and second order derivatives on a log *I*(*t*) scale. When this difference is constant across all values of *t*, then *I*(*t*) = exp(−*At*^2^ + *Bt* + *c*) holds and a Gaussian distribution is obtained, implying the epidemic is, in this sense, limited since as *t* → ∞, *I*(*t*) → 0.

In comparison, many large sample Gaussian statistical arguments, for example using the statistical score function [22], are based on a ratio, not a difference, of the first and second order derivatives of log(*I*(*t*)) This also underlies the Cramer-Rao bound or Fisher information bound on the sampling variation of maximum likelihood estimates.

Using continuous derivatives in relation to time, and assuming the numerator serves as an approximate second order change or acceleration term for log *I*(*t*), and the denominator an approximate first order change or derivative of log *I*(*t*), a continuous version of the BF condition can be expressed generally as:

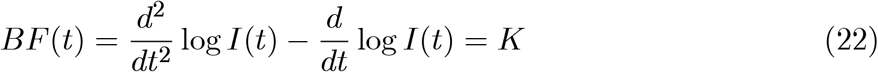

## 10 Pratt-Arrow Relative Risk

The BF mathematical condition above is similar to a generally defined measure of risk avoidance. The basic Pratt-Arrow measure of absolute relative risk ARA [13] can be defined as:

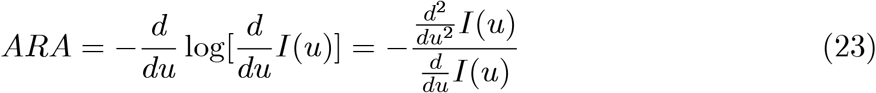

where *I*(*u*) is a function representing utility and *I*^′^ (*u*) > 0 for all *u*. While it is formally defined locally, and for individual behavior, if it is similar for all individuals, it can be viewed as a societal-level measure.

Individuals in an epidemic setting are subject to similar, mandated governmental restrictions, giving the ARA measure both individual and general social relevance. Note that the ARA measure, as a measure of utility, was not defined in order to meet an assumption of geometric progression, nor the onset of a Gaussian distribution.

As time affects the level of infection and thus relative risk and risk avoidance behavior for those who are not infected, this might also be more directly expressed as a function of infected counts through time, which are the primary sources of risk:

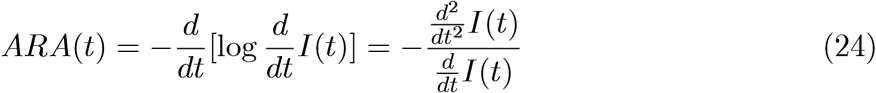

If the ARA measure of relative risk is constant, a condition similar in structure to the BF condition is obtained:

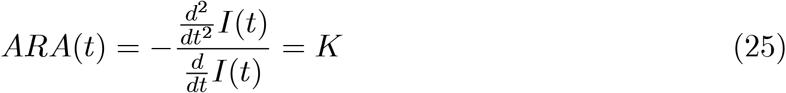

with risk avoidance behavior occuring where *K <* 1. Such a constant value for the ARA measure implies a stabilization in terms of risk avoidance behavior. Again, even though the ARA measure is defined at the individual level, the unique setting of shared pandemic restrictions for all individuals extends this restriction to society-level application.

## 11 Entropy and Maximum Entropy

The concept of entropy [24] can can be applied here to provide a comparative scale on which to examine the relative shapes of probability densities. Where the form of *I*(*t*) resulting from the BF condition is given by a formal probability distribution, the entropy of that distribution can provide a summary of the overall shape of the *I*(*t*) probability density. This can provide a scale on which the observed real-world histogram for *I*(*t*) can be compared to the Normal distribution.

The observed entropy of *I*(*t*) in general is defined here as:

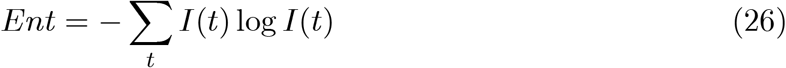

This can be obtained using the histogram of the observed relative frequency of infection cases.

If the BF condition holds, then:

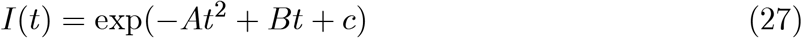

and the associated entropy is given by:

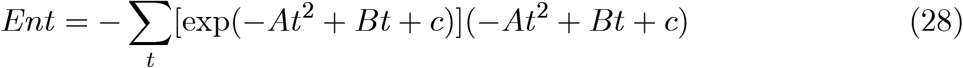

The concept of maximum entropy [25] is also relevant in obtaining the optimal form for *I*(*t*) under the BF restriction. Note that if *I*(*t*) has a parametric form, the observed value of the maxiumum likelihood estimate 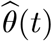 can be used to fit the distribution to the data as 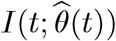. Indeed, the Normal distribution and uniform distributions can be interpreted as distributions possessing maximum entropy.

For example, in the specific case of the normal density, *N* (*µ, σ*^2^) entropy is given by:

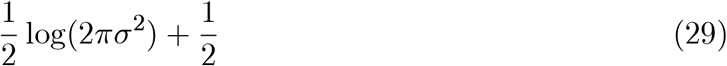

and can be estimated by:

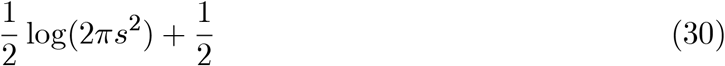

where *s*^2^ is the sample variance of the observed infection count data. This can be directly compared with the observed entropy from the observed histogram of *I*(*t*) values.

In the case of the exponential distribution, the distribution and entropy are given by:

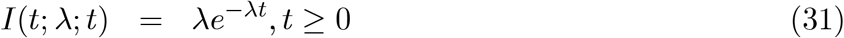

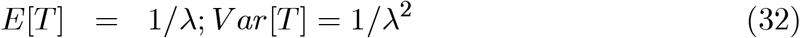

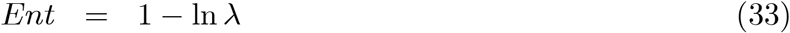

With the estimated entropy given by:

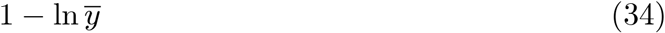

where 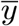 is the average number of infection cases over the time periods of relevance, for example monthly infections.

For distributions having unknown mean and variance parameters, the Normal distribution represents a distribution having maximum entropy [25]. For distributions having only an unknown mean parameter, the exponential distribution represents a maximum entropy distribution, and for simple discrete distributions, the uniform distribution represents a maximum entropy distribution.

## 12 Hazard Rate and Risk

The concept of risk, typically in the form of the onset of a fatal event, has been studied for many years in relation to insurance life tables and the related topic of survival analysis [26, 27]. A mathematical structure similar to the BF and Pratt-Arrow constructs can be applied in relation to risk defined as the instantaneous infection rate at time *t*, given non-infection up to time *t*.

The hazard rate (the risk of infection at any point in time, given non-infection up to that point) can be defined in terms of the survivor function *S*(*t*) = 1 − *F* (*t*), where *F* (*t*) is the cumulative probability density for *I*(*t*). Letting *T* denote the time of infection, the hazard rate can be defined in relation to the onset of infection:

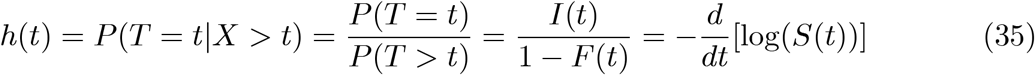

and a condition fairly similar to the BF condition can be defined, using the instantaneous infection rate, to reflect a constant, controlled hazard:

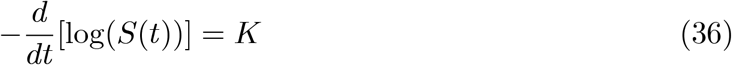

For example, with the exponential distribution, we have *I*(*t*) = *e*^−*λt*^ and *S*(*t*) = *e*^−*λt*^. Its hazard rate *h*(*t*) can be shown to equal λ, a constant. This implies an assumption of constant instantaneous risk or hazard if we assume the exponential distribution.

Note that in a mathematically similar context the BF condition uses the instantaneous first and second order derivatives of the incidence function log(*I*(*t*)), the hazard rate uses the cumulative probability density as a baseline and its first derivative relative to this baseline.

For example, in the case of the Weibull distribution for infection, *f* (*t*) = λ*γt*^*-γ*−l^ exp(−λ*t*^*-γ*^ ), its survivor function is given by *S*(*t*) = exp(−λ *t*^*-γ*^ ). It follows that its hazard function is given by 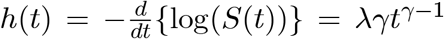. If *γ* > 1, the hazard or risk of infection is increasing over time and if *γ <* 1, it is decreasing. If *γ* = 1 the hazard is equal to λ, showing the exponential distribution to be a specific case of the Weibull distribution. This approach, along with Kaplan-Meier curves, can be used to compare time to infection across demographic groups.

## 13 Mathematical Similarities

It is interesting to examine epidemiologic phenomena as risk events at the macro or aggregate societal and governmental level, rather than focus on individual-level transmission dynamics as the onset of risk events often prompts common risk avoidance behavior in the entire unaffected human population.

The basic mathematical structure of risk and risk avoidance in the risk-related contexts considered here are similar. The BF condition and approach allows for the modeling of statistical properties that can be related to social utility based measures of risk avoidance, and also more standard measures of risk in the form of hazards, the hazard of becoming infected.

Consider the three basic representations of risk considered here:

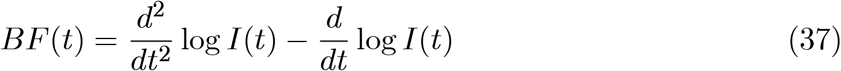

and

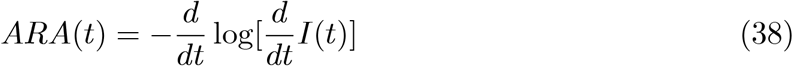

and

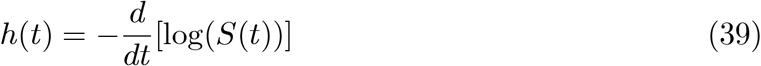

The differences in these measures of relative relate to changes in scale and the use of differences rather than ratios. The hazard rate *h*(*t*) measure uses changes in the cumulative probability density for *I*(*t*) on a log scale. The *BF* (*t*) measure uses the difference in second and first order derivatives with *I*(*t*) on a log scale. The *ARA*(*t*) measure uses a ratio of first and second order derivatives of *I*(*t*) without a log scale.

Note that these definitions of risk and related risk avoidance conditions come from very different areas of research and application, for example epidemiology, economics and actuarial science. In each, the study and modeling of perceived risk using the properties of the *I*(*t*) distribution has lead to similar mathematical structures of measurement and interpretation. These types of linkages may allow for the development of new areas of application for the various underlying model types, as well as new, extended versions of these models, implying greater flexibility in the choice of statistical model. This is the subject of ongoing work.

## 14 Discussion

Epidemics and pandemics are common in the history of societies. While they occur in real time, there is a period of rising incidence, sickness for the infected and rising peril for the non-infected. In such settings the increasing number of infected motivates risk avoidance behavior patterns on the part of society that remains healthy, often at the level of government restriction applying to all individuals. These patterns can be observed and modeled.

The BF restriction can be interpreted as the result of societal risk avoidance behavior in the presence of pandemic waves as individuals attempt to maximize risk avoidance in a given time period. It is mathematically similar in structure to the Pratt-Arrow ARA utility based risk avoidance behavioral measure. The condition also supports viewing the shape of *I*(*t*) in relation to entropy, with several of the probability distributions being interpretable as having maximum entropy.

Given the impact of an epidemic or pandemic on overall society, social risk and related utility functions are of interest. These will reflect the shape or entropy of the *I*(*t*) distribution, which reflects the overall infection rate. The shape of this function and the degree to which it differs from the applicable BF distribution or function, for example the Gaussian distribution reflecting maximum entropy, assuming the BF condition applies, may provide a measure of the extent to which the pandemic can be expected to continue.

The issue of optimizing behavior in the presence of widespread illness has been examined in the literature [28, 29]. These papers tend to modify the SIR or SEIR model to account for additional considerations related to maximizing utility at the individual level subject to illness related adjustments and additional restrictions to the SIR model.

To better understand the social processes affecting the possible limitation of an epidemic, it may be useful to examine the shape of the relative incidence distribution via entropy and formally model the onset of maximum entropy in the relative incidence function *I*(*t*), utilizing variables related to the measuring of social risk avoidance, and the formal structure of social utility functions. This macro-level or aggregate approach allows for the application of comparative statistical and probability related concepts to better understand how various risk based perspectives relate to each other.

For example, where the BF condition leads to a normal distribution, it can be interpreted as maximizing the entropy of the *I*(*t*) density. If the empirical probabilities for the histogram of observed *I*(*t*) values are available, then the entropy of the observed *I*(*t*) density can be obtained and may be compared to the entropy expected from the Gaussian distribution. Within a given time frame, this can be defined as Δ_*c*_, where:

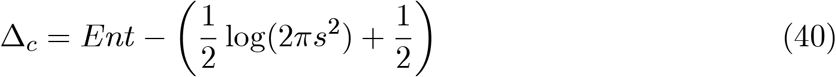

where *Ent* is the observed entropy of *I*(*t*). Values close to zero imply the BF condition is approximately true and the epidemic may be viewed as decreasing according to the BF condition over time. In settings where multiple Gaussian waves describe *I*(*t*), the entropy value can appropriately adjusted, defined basically as a weighted average.

The original work of Farr reflected consideration of epidemic waves through time. Spatial assessment of incidence counts can be assessed with the overall incidence being a mixture of locally defined *I* (*t*). If the BF condition for a given time period holds locally, for each spatial region or country considered, when social restrictions have imposed the condition, then the overall incidence function can be modeled as the weighted average of local distributions:

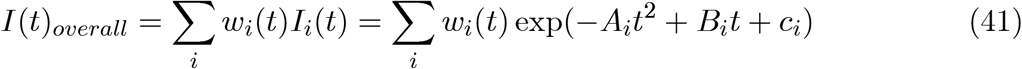

Practically, as a pandemic proceeds, the wavelength and frequency of the infection-waves decreases, often with telescoping weights. There is a societal learning effect as more waves are encountered. The imposition of social restrictions and resulting risk avoidance will have greater effects on later pandemic waves which will relatively weaken, subject to the caveat that the infectious agent does not genetically mutate. These will potentially differ by area allowing for different weighting schemes *w*_*i*_(*t*) and while they may overlap, it may be useful to examine such infection patterns within spatially defined sub-regions. Such waves have a long history in the epidemiologic and related gene wave literature [30]. This is the subject of future work.

## 15 Conclusions

The modeling of the frequency of new cases of infection in an epidemic within a given time period remains is of interest. Farr’s Law and the related Brownlee-Farr condition on infection counts implies a specific functional form for the related incidence function *I*(*t*) when the infectivity of an epidemic is decreasing. This form includes the Gaussian distribution and other distributions such as the exponential distribution. The Brownlee-Farr condition is shown here to correspond as a mathematical function to the Pratt-Arrow risk avoidance *ARA* measure, and may be interpreted as linking the Brownlee-Farr result to the concept of risk avoidance at the macro-societal level. This novel perspective also allows for the resulting BF condition to be compared to hazard function based measures of risk with notable similarities, reflecting different scales of measurement. This may allow for broader, more flexible approaches to the modeling of epidemics. As the shape of the *I*(*t*) distribution is an important aspect of understanding the onset of infection, the use of entropy and maximum entropy concepts may also be useful.

## Data Availability

All data produced in the present work are contained in the manuscript.

